# Anticoagulant vs. antiviral therapy, recovery and neurodamage in Long COVID: a real-world prospective cohort study

**DOI:** 10.1101/2025.02.14.25322178

**Authors:** Marc Jamoulle, Margaux Mignolet, Isabelle Meyts, Aurélie Ladang, Charles Nicaise, Johan Van Weyenbergh

## Abstract

Recent estimates consider that Long COVID affects 400 million people worldwide, of which <10% recover after two years. Predictive biomarkers and effective therapies are urgently needed for the clinical management of Long COVID. Neurological manifestations (poor memory, brain fog) are frequent, while neurodamage and neuroinflammatory markers UCHL1, GFAP and NFL were found to be increased in some but not all Long COVID cohorts. However, a direct link between two major Long COVID disease mechanisms, viral persistence and coagulation defects, and CNS damage is unclear. Therefore, we investigated the effect of combined anticoagulant (Asaflow + Clopidogrel) vs. antiviral (Paxlovid) treatment upon clinical outcome and neurodamage markers in Long COVID patients. Using multivariate logistic regression, we found that second-line antiviral treatment is the single best predictor of clinical recovery in a real-world Long COVID cohort (n=106) with long-term (220 person-years) follow-up. Our results support a putative role for viral persistence in Long COVID pathogenesis but also indicate a large therapeutic window for Paxlovid and other antivirals. A rapid decline in neurodamage markers (GFAP/NFL) upon antiviral treatment suggests CNS damage in Long COVID might be therapeutically targeted and should be considered in ongoing and future clinical trials.

---

Recent estimates consider that Long COVID affects 400 million people worldwide, of which <10% recover after two years^1^. Predictive biomarkers and effective therapies are urgently needed for the clinical management of Long COVID^2^. Neurological manifestations (poor memory, brain fog) are frequent^3^, while neurodamage and neuroinflammatory markers UCHL1, GFAP and NFL were found to be increased^4^ in some but not all Long COVID cohorts. However, a direct link between viral persistence and coagulation defects, two proposed Long COVID disease mechanisms^5,6^, and CNS damage is unclear. Therefore, we investigated the effect of anticoagulant and antiviral treatment upon clinical outcome and neurodamage markers in Long COVID patients.

In a real-word cohort with long-term follow-up (220 person-years, n=106), we measured both patient-(COOP chart score ranging 0-30) and clinician-reported (Long Covid Grade ranging 0-3) outcomes following first-line anticoagulant treatment (Asaflow + Clopidogrel), and optional second-line 15-day antiviral treatment with Paxlovid (Nirmatrelvir/Ritonavir). Out of 106 patients, 73 received anticoagulant prescription, 17 received antiviral prescription, while 16 received neither. Patient-reported improvement shows antiviral treatment as independent predictor (OR 6.18, 95%CI 1.35-34.13, p=0.024), while age, female sex, the number of vaccine doses, disease severity, COVID wave/variant, and anticoagulant treatment were not. No significant predictors were observed for clinician-reported clinical improvement. Antiviral (OR 4.57, 95%CI 1.19-20.74) but not anticoagulant (OR 0.35, 95%CI 0.055-1.92) treatment was a significant predictor of strictly or broadly defined (patient- and/or clinician-measured) clinical improvement in multivariate logistic regression, independent of age, sex, vaccine doses, disease severity, comorbidities, and COVID wave/variant (Fig. 1A-B).

**Figure 1:**
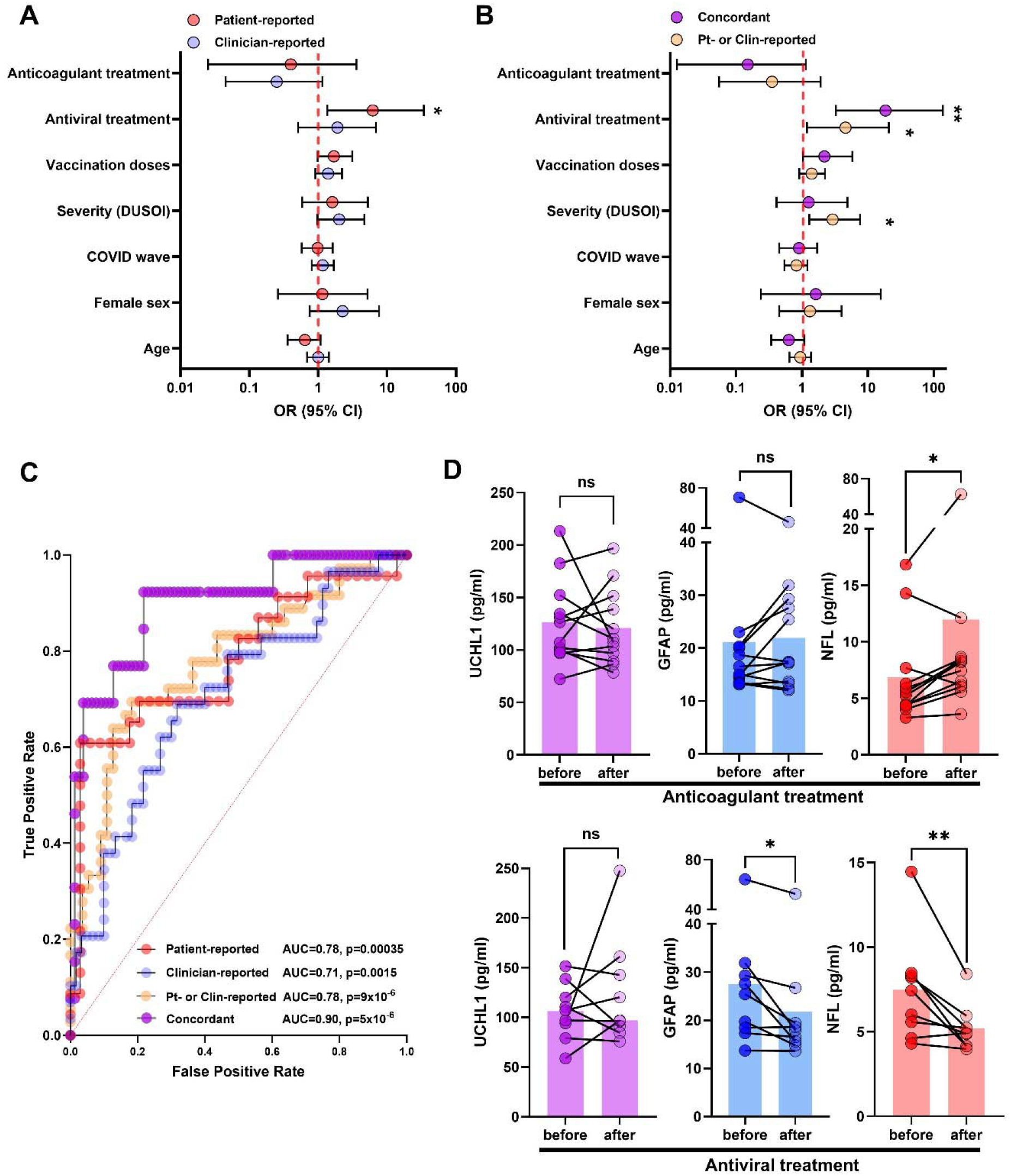
Antiviral therapy is an independent predictor of concordant clinician- and patient-reported clinical improvement, and decreases systemic markers of neurodamage and neuroinflammation in Long COVID patients. A) Forest plot showing multivariate logistic regression models for patient- and clinician-reported clinical improvement, as measured by a decrease in COOP chart score (see Supplementary fig. S1) and clinical grade (rated on a scale from 0-3), respectively. B) Forest plot showing multivariate logistic regression models for broadly defined clinical improvement (patient or clinician-reported) and strictly defined (concordant patient- and clinician-reported clinical improvement), as measured by a decrease in both COOP chart score and clinical grade. C) ROC curve analysis shows improved prediction of strictly defined (concordant patient- and clinician-reported, AUC=0.90 95%CI 080-0.99) over broadly defined (patient- or clinician-reported, AUC=0.78, 95%CI 0.68-0.88), patient-reported (AUC=0.78, 95%CI 0.65-0.91) or clinician-reported (AUC=0.71, 95%CI 0.59-0.82) clinical improvement. D) In a subset of patients with longitudinal sampling (n=12 for anticoagulant treatment (top panel), n=9 for antiviral treatment, (lower panel)), markers of neurodamage/neuroinflammation were unchanged (UCHL1, GFAP, p>0.05) or increased (NFL, p=0.017) after anticoagulant treatment. GFAP (p=0.012) and NFL (p=0.0078) significantly decreased after antiviral treatment. (*p<0.05, **p<0.01, Wilcoxon Matched Pairs test)

In a first sensitivity analysis, the addition of comorbidities as an additional parameter to the regression models did not significantly change the odds ratios or ROC curves for any of the four models (data not shown). Secondly, patients with antiviral treatment were removed (n=17), revealing no independent predictors in multivariate logistic regression and no significant AUC in ROC curve analysis (detailed in Supplementary Table 2).

Predictive power was confirmed by ROC curves (Fig. 1C), with improved prediction of strictly defined (concordant patient- and clinician-reported, AUC=0.90 95%CI 080-0.99) over broadly defined (patient- or clinician-reported, AUC=0.78, 95%CI 0.68-0.88), patient-reported (AUC=0.78, 95%CI 0.65-0.91) or clinician-reported (AUC=0.71, 95%CI 0.59-0.82) clinical improvement.

In a subset of patients with longitudinal sampling (Fig. 1D), serum markers of neurodamage were unchanged (UCHL1/GFAP) or increased (NFL, p=0.017, n=12) after anticoagulant treatment, while both GFAP (p=0.012, n=9) and NFL (p=0.0078, n=9) significantly decreased after antiviral treatment. Although sample numbers were small, a homogeneous and fast (15-day) response in neurodamage (GFAP/NFL) markers after antiviral treatment suggests a direct link between an uncharted viral reservoir and CNS damages. Limitations of the study include the small sample size, as well as missing data, the latter being inherent to real-world cohorts. Strengths of the study include a general practice-based real world cohort with complete electronic health records, uniform first and second-line treatment options, and the use of validated scales for both patient-reported and clinician-reported outcomes (see Supplementary Data).

In conclusion, second-line antiviral treatment is the single best predictor of clinical recovery in a real-world Long COVID cohort, supporting a putative role for viral persistence in Long Covid. A rapid decline in neurodamage markers upon antiviral treatment suggests CNS damage in Long COVID might be therapeutically targeted and should be considered in ongoing and future clinical trials.

## Data Availability

All data produced in the present study are available upon reasonable request to the authors.

## Disclosure of all financial associations or other possible conflicts of interest

JVW received funding from Research Foundation Flanders (FWO) grants G0A0621N and G065421N, PolyBio Research Foundation, and LCAP/LCF crowdfunding. MJ received a grant from King Baudouin Foundation (2022-J51708200-F001). IM received grants from UNDINE Horizon Europe, CSL Behring and Octapharma. IM received consulting/speaker fees from Boehringer Ingelheim, Takeda, and CSL Behring; JVW received a speaker fee from Pfizer. The remaining authors declare no competing interests.

