## Supplementary Data for "Anticoagulant vs. antiviral therapy, recovery and neurodamage in Long COVID: a real-world prospective cohort study"

This study was approved by the Ethical committees of University Hospitals Leuven (UZ Leuven, Belgium) and University Hospital CHU ULC (Namur, Belgium), all patients have provided written informed consent.

Demographic and clinical data from all 106 Long COVID patients are summarized in Supplementary Table 1. A diagnosis of Long COVID was made according to WHO criteria^1^: “defined as the continuation or development of new symptoms 3 months after the initial SARS-CoV-2 infection, with these symptoms lasting for at least 2 months with no other explanation.” Patient-reported outcomes were measured using validated COOP/WONCA charts^2^ (see Supplementary Figure 1), clinician-reported outcomes were measured using DUSOI (Duke University Severity Of Illness) score^3^, and Long COVID grade (score from 0-3), as previously described^4-5^.

Acute COVID-19 infection and comorbidities were documented for all patients using the national database of electronic health records, which is centralized and linked to a single general practician per patient in the Belgian healthcare system (access granted to first author MJ for all patients).

First-line dual anticoagulant treatment (Asaflow 80 mg/day + Clopidogrel 75 mg/day) was prescribed to all patients with severe disease (clinician-reported Long COVID grade 2 or 3), while second-line antiviral treatment (Paxlovid, Nirmatrelvir 300 mg and Ritonavir 100 mg, twice daily for 15 days) was offered as an opt-in choice for patients, at which time the anticoagulant treatment was discontinued. Serum markers were quantified by sensitive SIMOA assays (GFAP and UCHL1 by Alinity® Abbott, NFL by Lumipulse® G from Fujirebio) and reported in pg/ml.

Statistical tests performed with GraphPad Prism and XL-STAT software included multivariable logistic regression, normality testing (Shapiro-Wilk and Kolmogorov-Smirnov tests) guided subsequent parametric or non-parametric (Wilcoxon matched-pairs signed rank test, Spearman correlation) analysis, all two-tailed. Missing data were not imputed, thus the final multivariable logistic regression (Fig. 1A-B) was performed on 91 out of 106 patients.

**Supplementary Table 1: Clinical and demographic data for Long COVID patients (n=106)**

|  | **Antiviral treatment (n=17)** | | **Anticoagulant treatment (n=73)** | | **No treatment (n=16)** | |
| --- | --- | --- | --- | --- | --- | --- |
|  | **Number/Mean** | **SD** | **Number/Mean** | **SD** | **Number/Mean** | **SD** |
| **Age** | 46.9 | 13.7 | 46.6 | 13.1 | 41.1 | 19.0 |
| **Sex** | 12F/5M |  | 47F/16M |  | 10F/6M |  |
| **WT/D614G** | 7 |  | 37 |  | 8 |  |
| **Alpha** | 3 |  | 8 |  | 1 |  |
| **Delta** | 2 |  | 7 |  | 3 |  |
| **Omicron** | 5 |  | 20 |  | 3 |  |
| **Number of comorbidities** | 3.9 | 4.1 | 4.3 | 2.9 | 2.3 | 1.5 |
| **Global Severity (DUSOI)** | 3.6 | 0.5 | 3.4 | 0.7 | 1.9 | 0.8 |
| **Time since acute Covid (months)** | 24.5 | 10.7 | 23.8 | 11.1 | 19.5 | 14.7 |
| **Vaccination doses** | 2.4 | 1.3 | 2.0 | 1.3 | 1.7 | 1.3 |
| **First COOP Chart total** | 24.3 | 2.2 | 22.1 | 5.9 | 17.8 | 5.7 |
| **Last COOP Chart total** | 21.9 | 4.7 | 22.4 | 4.4 | 16.5 | 7.0 |
| **First LC grade** | 3.0 | 0.0 | 2.6 | 0.5 | 1.3 | 0.7 |
| **Last LC Grade** | 2.4 | 0.8 | 2.4 | 0.7 | 0.8 | 0.6 |

*Data represent the number of patients (sex and SARS-CoV-2 variant) or the mean and standard deviation (SD). Comorbidities were identified in Belgian national registry electronic health records and classified according to ICPC-2^6^. DUSOI score (clinician-reported) ranges from 1-5, Long COVID grade ranges from 0-3 (clinician-reported), COOP total score (patient-reported) ranges from 6-30, higher scores correspond to worse clinical status for all scales.*

**Supplementary Table 2: Sensitivity analysis (removing patients with antiviral treatment n=17) Multivariable logistic regression models for classification of Long COVID patients with and without clinical improvement (n=65)**

| **Odds ratios** | **Variable** | **Estimate** | **95% CI** | **P value** |
| --- | --- | --- | --- | --- |
| β1 | Age | 1.00 | 0.94 - 1.07 | 0.92 |
| β2 | Sex | 1.01 | 0.21 - 4.60 | 0.99 |
| β3 | Comorbidities | 1.04 | 0.77 - 1.35 | 0.80 |
| β4 | COVID wave | 0.97 | 0.52 - 1.70 | 0.91 |
| β5 | Severity (DUSOI) | 2.86 | 0.93 - 13.22 | 0.13 |
| β6 | Vaccination doses | 1.18 | 0.64 - 2.34 | 0.60 |
| β7 | Anticoagulant treatment | 0.47 | 0.054 - 4.86 | 0.50 |
| **Area under the ROC curve** |  |  |  |  |
| Area | 0.68 |  |  |  |
| Std. Error | 0.08 |  |  |  |
| 95% confidence interval | 0.53 to 0.84 |  |  |  |
| P value | 0.057 |  |  |  |

**Supplementary Figure 1: Visual chart (COOP/WONCA) used to quantify patient-reported outcomes in six dimensions (physical fitness, feelings, daily activities, social activities, change in health, overall health), each on a visual scale from 1 to 5 (higher scores means worse status).**

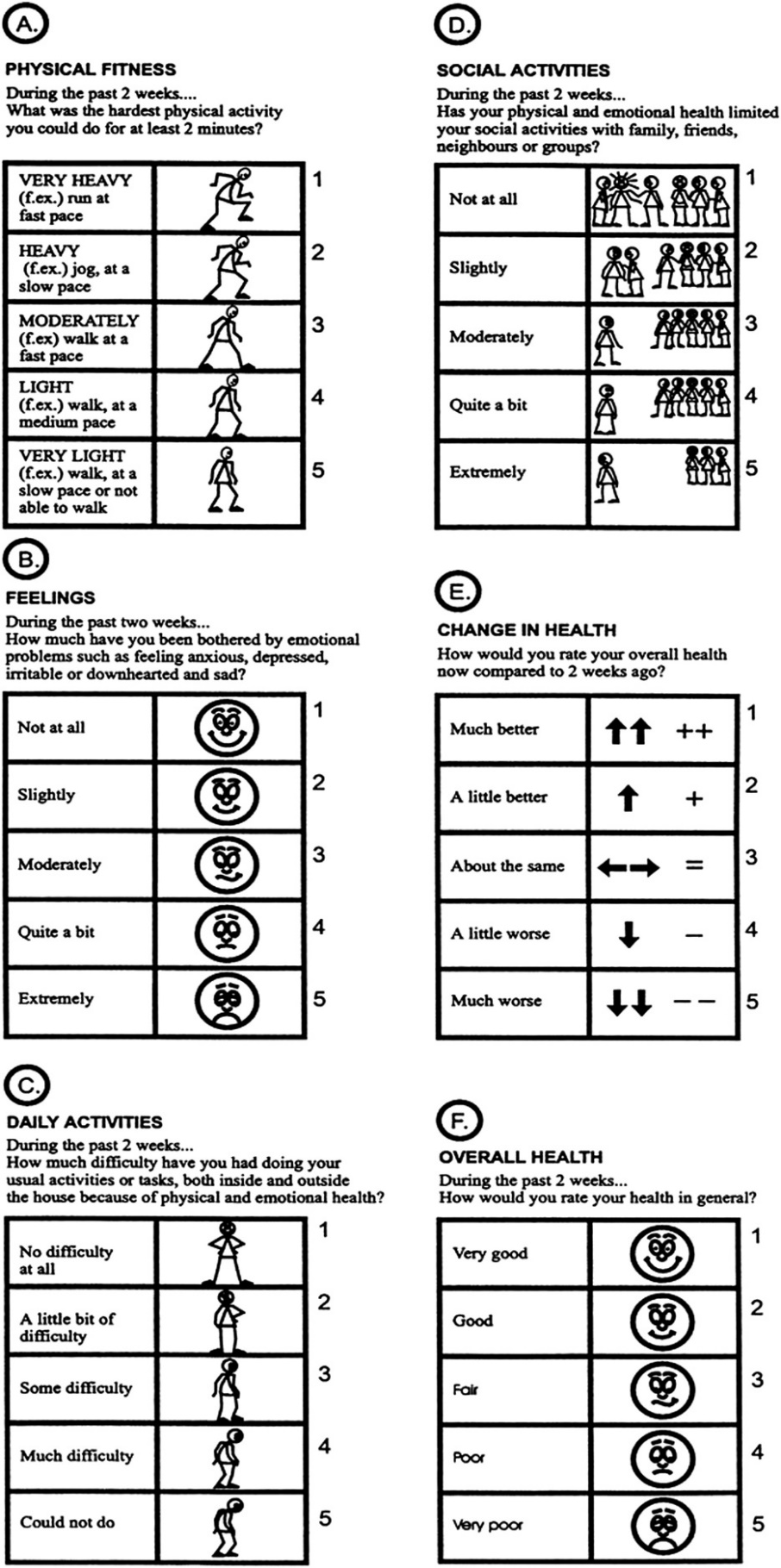
